# AI-Powered Social Robots for Addressing Loneliness: A Systematic Review Protocol

**DOI:** 10.1101/2025.06.19.25329905

**Authors:** Ravi Shankar, Fiona Devi, Xu Qian

## Abstract

Loneliness is a significant public health concern that affects millions of people worldwide, particularly older adults. With advancements in artificial intelligence (AI) and robotics, researchers are exploring the potential of AI-driven social robots as a means to alleviate loneliness and provide companionship. This systematic review protocol aims to identify, evaluate, and synthesize existing literature on the effectiveness of AI-driven social robots in combating loneliness among older adults. The review will follow the Preferred Reporting Items for Systematic Reviews and Meta-Analyses (PRISMA) guidelines and search multiple electronic databases from their inception to June 2025, including PubMed, Web of Science, Embase, CINAHL, MEDLINE, The Cochrane Library, PsycINFO, and Scopus. Eligible studies will be those that involve AI-driven social robots designed to alleviate loneliness in older adults, with outcomes related to loneliness, social isolation, or quality of life. Two reviewers will independently screen studies using Covidence software, extract data, and assess the risk of bias using the Cochrane Risk of Bias Tool. A narrative synthesis of the findings will be conducted, with meta-analysis performed if the data allows. This review will provide valuable insights into the current state of research on AI-driven social robots for combating loneliness and guide future research and development in this area.

## 1. Introduction

Loneliness is a subjective and distressing feeling arising from perceived deficiencies in the quantity or quality of one’s social relationships [1]. It is a multifaceted construct that encompasses both emotional loneliness, which refers to the absence of an intimate attachment figure, and social loneliness, which refers to the lack of a broader group of contacts or an engaging social network [2]. Loneliness is distinct from objective social isolation, as it is possible to be socially isolated without feeling lonely, or to feel lonely despite having social connections [3].

The prevalence of loneliness has been recognized as a significant public health issue, with studies suggesting that it affects a substantial proportion of the population across different age groups and cultures [4]. However, older adults are particularly vulnerable to experiencing loneliness, as they often face multiple risk factors such as the loss of loved ones, retirement, decreased mobility, and chronic health conditions [5]. A meta-analysis by Ong et al. [6] found that the prevalence of loneliness among older adults ranged from 5% to 43% across various studies, with a pooled prevalence of 28.5%.

The negative impact of loneliness on physical and mental health has been well-documented in the literature. Loneliness has been associated with an increased risk of all-cause mortality [7], cardiovascular disease [8], cognitive decline and dementia [9], depression [10], and suicide [11]. Moreover, loneliness has been linked to unhealthy behaviors such as smoking, physical inactivity, and poor sleep [12], which can further contribute to adverse health outcomes. Given the significant burden of loneliness on individuals and society, there is a pressing need for effective interventions to alleviate loneliness and promote social connectedness among older adults.

In recent years, there has been growing interest in the potential of AI-driven social robots as a novel approach to combating loneliness in older adults. Social robots are defined as autonomous or semi-autonomous robots that interact and communicate with humans by following social behaviors and rules [13]. With advances in AI technologies such as natural language processing, emotion recognition, and machine learning, social robots are becoming increasingly sophisticated in their ability to engage in social interactions and provide companionship [14].

Several studies have explored the use of social robots in various settings, including homes, nursing homes, and hospitals, with promising results. For example, a randomized controlled trial by Robinson et al. [15] found that the use of a companion robot named Paro, which resembles a baby harp seal, significantly reduced loneliness and improved mood among residents in a nursing home. Another study by Moyle et al. [16] reported that the use of a humanoid robot named Pepper led to increased engagement and social interaction among older adults with dementia in a residential care facility.

However, despite the growing body of research on social robots and loneliness, there remains a lack of systematic evidence on their effectiveness and optimal implementation. Previous reviews on this topic have been limited by their narrow focus on specific types of robots or settings [17], or by their inclusion of studies with diverse populations and outcomes [18]. Moreover, the rapid advancement of AI technologies and the increasing availability of social robots warrant an updated review of the most recent evidence.

This systematic review protocol aims to address these gaps by providing a comprehensive and up-to-date synthesis of the literature on AI-driven social robots for combating loneliness in older adults. By following a rigorous and transparent methodology, this review will offer valuable insights into the current state of the evidence, identify key knowledge gaps, and inform future research and practice in this emerging field.

The overarching question that this systematic review seeks to answer is: What is the effectiveness of AI-driven social robots in reducing loneliness and improving related outcomes among older adults, compared to usual care or other interventions? To further guide the review, several sub-questions will be explored. First, what are the characteristics and components of AI-driven social robots that have been used to combat loneliness in older adults? Second, what are the effects of these robots on outcomes such as loneliness, social isolation, social support, quality of life, and other relevant measures in older adults? Third, are there differences in effectiveness based on participant characteristics (e.g., age, gender, living arrangement), intervention characteristics (e.g., type of robot, duration, frequency), or study characteristics (e.g., study design, setting)? Fourth, what are the experiences and perspectives of older adults, caregivers, and healthcare professionals regarding the acceptability, feasibility, and usability of AI-driven social robots? Finally, the review will examine any potential adverse effects or unintended consequences associated with the use of such robots in this population.

## 2. Methods

### 2.1. Review design

This systematic review will be conducted in accordance with the Preferred Reporting Items for Systematic Reviews and Meta-Analyses (PRISMA) statement [19] and the Cochrane Handbook for Systematic Reviews of Interventions [20]. The protocol for this review has been registered with the International Prospective Register of Systematic Reviews (PROSPERO) (registration number: CRD42025641046).

### 2.2. Eligibility criteria

#### 2.2.1. Types of studies

This review will include primary studies employing various research designs, including randomized controlled trials (RCTs), cluster RCTs, quasi-experimental studies (such as non-randomized controlled trials, controlled before-and-after studies, and interrupted time series), cohort studies, case-control studies, cross-sectional studies, and qualitative studies. Conference abstracts, editorials, commentaries, and opinion pieces will be excluded. While systematic reviews and meta-analyses will not be included in the analysis, their reference lists will be screened to identify potentially relevant primary studies.

#### 2.2.2. Types of participants

This review will include studies involving older adults aged 60 years and above who are at risk of or experiencing loneliness. Studies conducted in any setting (e.g., community, nursing homes, hospitals) and in any geographical location will be eligible.

Studies that focus on specific sub-populations of older adults (e.g., those with dementia, depression, or other medical conditions) will be included if loneliness or related outcomes are reported. Studies involving participants of all ages will be included if data for older adults can be extracted separately.

#### 2.2.3. Types of interventions

This review will include studies that evaluate the use of AI-driven social robots as an intervention for combating loneliness in older adults. For the purpose of this review, AI-driven social robots are defined as robots that are designed to interact socially with humans and that incorporate AI technologies such as natural language processing, speech recognition, computer vision, and machine learning to enable adaptive and personalized interactions.

The AI-driven social robots can take various forms (e.g., humanoid, animal-like, or abstract), and can be used for different purposes (e.g., companionship, therapy, education, or entertainment). Studies that involve the use of robots without AI capabilities (e.g., simple telepresence robots) or robots that are not specifically designed for social interaction (e.g., rehabilitation robots) will be excluded.

Eligible interventions can be delivered in different settings (e.g., home, nursing home, hospital), and can be used alone or in combination with other interventions (e.g., psychosocial interventions, medication). Studies that compare the effectiveness of different types of AI-driven social robots will also be included.

#### 2.2.4. Types of comparators

This review will include studies that compare the effectiveness of AI-driven social robots to one of the following comparators: usual care (such as standard social activities or programs), no intervention or waitlist control, and other interventions aimed at reducing loneliness (e.g., psychosocial interventions, pet therapy, or human companionship). Studies that do not include a comparator group will be excluded from the review.

#### 2.2.5. Types of outcomes

The primary outcome of interest for this review is loneliness, assessed using validated measurement tools such as the UCLA Loneliness Scale, the De Jong Gierveld Loneliness Scale, or the Campaign to End Loneliness Measurement Tool. Secondary outcomes include social isolation, which may be measured through objective indicators such as social network size, frequency of social contacts, or participation in social activities; and social support, assessed using instruments like the Multidimensional Scale of Perceived Social Support or the Lubben Social Network Scale. Quality of life will also be examined, using generic or disease-specific tools such as the World Health Organization Quality of Life (WHOQOL) scale or the EuroQol Five-Dimension (EQ-5D) scale. Additional secondary outcomes include depressive symptoms (e.g., measured by the Geriatric Depression Scale or the Patient Health Questionnaire-9), anxiety symptoms (e.g., measured by the Generalized Anxiety Disorder-7 scale or the State-Trait Anxiety Inventory), and cognitive function (e.g., assessed using the Mini-Mental State Examination or the Montreal Cognitive Assessment). The review will also consider adverse events, including any reported physical or psychological harm, unintended consequences, or technical issues related to the use of AI-driven social robots. Finally, qualitative outcomes—such as the experiences and perspectives of older adults, caregivers, and healthcare professionals—will be included when reported in the eligible studies.

### 2.3. Search strategy

A comprehensive search strategy will be developed in consultation with a health sciences librarian to identify all relevant studies. The following electronic databases will be searched from their inception to June 2025: PubMed, Web of Science, Embase, CINAHL, MEDLINE, The Cochrane Library, PsycINFO, and Scopus.

The search strategy will use a combination of keywords and controlled vocabulary terms (e.g., MeSH terms) related to the following concepts: (1) older adults, (2) loneliness or social isolation, (3) social robots, and (4) artificial intelligence. An example search string for PubMed is provided in Table 1.

**Table 1.** Search strategy for PubMed database.

| Search Component | Search Terms |
| --- | --- |
| **Population** | ("aged"[MeSH Terms] OR "older adult*" OR "elderly" OR "senior*" OR "geriatric*" OR "aging" OR "ageing" OR "60 years" OR "65 years" OR "70 years") |
| **Condition** | ("loneliness"[MeSH Terms] OR "lonely" OR "social isolation"[MeSH Terms] OR "social isolat*" OR "social disconnect*" OR "social withdraw*" OR "emotional loneliness" OR "social loneliness") |
| **Intervention** | ("robotics"[MeSH Terms] OR "robot*" OR "social robot*" OR "companion robot*" OR "assistive robot*" OR "interactive robot*" OR "therapeutic robot*" OR "humanoid robot*") |
| **AI Component** | ("artificial intelligence"[MeSH Terms] OR "AI" OR "machine learning" OR "natural language processing" OR "computer vision" OR "speech recognition" OR "adaptive interact*" OR "intelligent system*" OR |
|  | "conversational agent*") |
| **Final Search String** | (#1 Population) AND (#2 Condition) AND (#3 Intervention) AND (#4 AI Component) |
| **Filters Applied** | None (no language, date, or publication type restrictions will be applied) |

Asterisks (*) in the above table represent truncation to capture word variations, and MeSH terms will be exploded to include all subheadings. This search strategy will be adapted for each database using appropriate controlled vocabulary terms, such as EMTREE terms for Embase and CINAHL headings for CINAHL. Boolean operators (AND, OR) will be adjusted according to database-specific syntax requirements to ensure optimal retrieval of relevant studies across all searched databases.

In addition to the electronic database search, several other sources will be explored to identify additional relevant studies. These include the reference lists of included studies and relevant reviews, clinical trial registries such as ClinicalTrials.gov and the WHO International Clinical Trials Registry Platform, and grey literature databases like OpenGrey and ProQuest Dissertations & Theses Global. Conference proceedings and abstracts from events such as the International Conference on Social Robotics and the IEEE International Conference on Robotics and Automation will also be reviewed. Furthermore, websites of relevant organizations and research groups, including the IEEE Robotics and Automation Society and the Association for the Advancement of Artificial Intelligence, will be examined. Experts in the fields of social robotics and gerontology will be contacted to identify any additional studies or unpublished data.

No language or publication date restrictions will be applied to the search. Studies published in languages other than English will be translated using Google Translate or by a native speaker if feasible.

### 2.4. Study selection

The study selection process will be conducted in two stages using Covidence, a web-based software platform for managing systematic reviews [34].

In the first stage, two reviewers will independently screen the titles and abstracts of all records retrieved from the search against the eligibility criteria. Records that clearly do not meet the criteria will be excluded, while those that appear potentially eligible or unclear will be retained for full-text review. Any discrepancies between the reviewers will be resolved through discussion or by consulting a third reviewer if necessary.

In the second stage, the full-text articles of the remaining records will be retrieved and independently assessed by two reviewers against the eligibility criteria. Reasons for exclusion at this stage will be recorded and reported in the final review. Any disagreements between the reviewers will be resolved through discussion or by consulting a third reviewer if necessary.

The study selection process will be documented using a PRISMA flow diagram [19], which will show the number of records identified, included, and excluded at each stage of the review.

### 2.5. Data extraction

Data from the included studies will be extracted using a standardized data extraction form in Covidence. The form will be pilot-tested on a sample of studies and refined as necessary. Two reviewers will independently extract data from each included study, and any discrepancies will be resolved through discussion or by consulting a third reviewer if necessary.

The following data will be extracted from each included study:

- Study characteristics: authors, year of publication, country, study design, sample size, setting, duration of follow-up
- Participant characteristics: age, gender, ethnicity, education level, living arrangement, health status, level of loneliness or social isolation at baseline
- Intervention characteristics: type of AI-driven social robot, description of AI technologies used, purpose of the robot, duration and frequency of use, any co-interventions
- Comparator characteristics: type of comparator, description of usual care or other interventions, duration and frequency of use
- Outcomes: loneliness, social isolation, social support, quality of life, depressive symptoms, anxiety symptoms, cognitive function, adverse events, and any other relevant outcomes reported in the study
- Qualitative data: experiences and perspectives of participants, caregivers, and healthcare professionals regarding the acceptability, feasibility, and usability of the intervention
- Study funding sources and conflicts of interest

If any relevant data are missing or unclear in the published reports, the study authors will be contacted via email to request additional information.

### 2.6. Risk of bias assessment

The risk of bias in the included studies will be assessed using appropriate tools tailored to the study design. For randomized controlled trials (RCTs), the Cochrane Risk of Bias 2 (RoB 2) tool will be employed, which evaluates five domains: the randomization process, deviations from intended interventions, missing outcome data, measurement of the outcome, and selection of the reported result. For non-randomized studies, the Risk Of Bias In Non-randomized Studies of Interventions (ROBINS-I) tool will be used, assessing bias due to confounding, participant selection, classification of interventions, deviations from intended interventions, missing data, outcome measurement, and selection of the reported result. Each domain in these tools will be rated as having low, moderate, serious, or critical risk of bias based on the respective tool’s criteria. For qualitative studies, the Critical Appraisal Skills Programme (CASP) Qualitative Checklist will be applied. This checklist examines ten domains: a clear statement of aims, appropriate methodology, research design, recruitment strategy, and data collection, consideration of the researcher-participant relationship, ethical issues, rigor of data analysis, clarity of findings, and the overall value of the research.

Two reviewers will independently assess the risk of bias in each included study, and any disagreements will be resolved through discussion or by consulting a third reviewer if necessary. The results of the risk of bias assessment will be reported in the final review using appropriate tables or figures.

### 2.7. Data synthesis

If the included studies are sufficiently homogeneous in terms of participants, interventions, comparators, and outcomes, a meta-analysis will be conducted to quantitatively synthesize the results. For continuous outcomes, the mean difference (MD) or standardized mean difference (SMD) with 95% confidence intervals (CIs) will be calculated, depending on whether the same or different measurement scales are used across studies. For dichotomous outcomes, the risk ratio (RR) or odds ratio (OR) with 95% CIs will be calculated. Heterogeneity between studies will be assessed using the I² statistic and the Chi² test, with I² values of 25%, 50%, and 75% considered as low, moderate, and high heterogeneity, respectively [38]. If significant heterogeneity is detected (I² > 50% or P < 0.1), a random-effects model will be used for meta-analysis; otherwise, a fixed-effect model will be used.

Subgroup analyses will be performed to explore potential sources of heterogeneity and to examine the effectiveness of AI-driven social robots across different subpopulations or settings, provided that sufficient data are available. Potential subgroups of interest include age groups (e.g., 60–69, 70–79, 80+ years), gender (male vs. female), living arrangement (community-dwelling vs. residential care), health status (healthy vs. frail or with specific medical conditions), type of robot (humanoid vs. animal-like vs. abstract), duration of intervention (short-term vs. long-term), and study design (RCTs vs. non-randomized studies).

Sensitivity analyses will be performed to assess the robustness of the meta-analysis results by excluding studies with high risk of bias or by using alternative methods for handling missing data.

If meta-analysis is not feasible due to significant heterogeneity or insufficient data, a narrative synthesis will be conducted to summarize the findings of the included studies. The narrative synthesis will be structured around the review questions and will address several key aspects, including the direction and magnitude of effects, the consistency of effects across studies, the strength of the evidence based on study quality and the precision of effect estimates, and the applicability of the evidence to various populations and settings. Tables and figures will be used to present the characteristics and findings of the included studies in a clear and concise manner.

### 2.8. Grading the quality of evidence

The quality of evidence for each outcome will be assessed using the Grading of Recommendations Assessment, Development and Evaluation (GRADE) approach. This method evaluates evidence based on several domains: risk of bias, inconsistency, indirectness, imprecision, publication bias, large effect size, dose-response gradient, and residual confounding. Based on these criteria, the quality of evidence will be rated as high, moderate, low, or very low. Two reviewers will make these judgments independently, and any disagreements will be resolved through discussion or, if necessary, by consulting a third reviewer. The findings of the GRADE assessment will be presented in a summary of findings table, which will include the number of participants and studies, effect estimates with 95% confidence intervals, the assigned quality of evidence, and explanatory comments or footnotes supporting the judgments.

### 2.9. Reporting bias assessment

If meta-analysis is conducted, publication bias will be assessed using funnel plots and statistical tests such as Egger’s test [40] or Begg’s test [41], if there are at least 10 studies included in the meta-analysis. Asymmetry in the funnel plot or significant results from the statistical tests may indicate the presence of publication bias.

In addition, selective reporting bias will be assessed by comparing the outcomes reported in the published articles with those listed in the study protocols or trial registries, if available. Discrepancies between the reported and planned outcomes may suggest selective reporting bias.

The results of the reporting bias assessment will be considered in the interpretation and discussion of the review findings.

## 3. Discussion

This systematic review protocol outlines a comprehensive and rigorous plan to synthesize the available evidence on the effectiveness of AI-driven social robots in combating loneliness among older adults. By following a transparent and systematic approach, this review aims to provide a reliable and up-to-date assessment of the state of the evidence and to identify key knowledge gaps and future research priorities.

The strengths of this protocol include the use of a broad and inclusive search strategy, the involvement of multiple databases and grey literature sources, and the adherence to established guidelines and tools for conducting and reporting systematic reviews. The inclusion of both quantitative and qualitative studies will allow for a more comprehensive understanding of the effectiveness, acceptability, and feasibility of AI-driven social robots from different perspectives.

However, there are also several limitations and challenges that need to be acknowledged. First, the rapid pace of development in the field of social robotics means that the evidence base is likely to evolve quickly, and the findings of this review may need to be updated regularly. Second, the heterogeneity of the included studies in terms of robot types, intervention designs, and outcome measures may limit the ability to draw firm conclusions and to conduct meaningful meta-analyses. Third, the inclusion of non-randomized studies may increase the risk of bias and confounding, which will need to be carefully assessed and considered in the interpretation of the results.

Despite these limitations, this systematic review has the potential to make important contributions to the field of gerontechnology and to inform the development and implementation of AI-driven social robots for combating loneliness in older adults. By synthesizing the available evidence, this review can help to identify the most promising approaches and the key factors that influence their effectiveness, as well as the potential barriers and facilitators to their adoption and scalability.

The findings of this review can also have important implications for policy and practice. For example, they can inform the design of public health interventions and the allocation of resources for supporting the social and emotional well-being of older adults. They can also guide the development of guidelines and standards for the ethical and responsible use of AI-driven social robots in healthcare and social care settings.

In addition, this review can contribute to the broader discourse on the role of technology in addressing the challenges of population aging and the importance of considering the social and psychological dimensions of health and well-being in the design and evaluation of technological interventions.

In conclusion, this systematic review protocol presents a timely and important initiative to advance our understanding of the potential of AI-driven social robots in combating loneliness among older adults. By providing a comprehensive and rigorous synthesis of the available evidence, this review can inform the development of more effective and acceptable interventions and support the well-being and quality of life of older adults in an increasingly digital world.

## Data Availability

All data produced in the present work are contained in the manuscript

## References

[1] Peplau LA, Perlman D. Perspectives on loneliness. In: Peplau LA, Perlman D, editors. Loneliness: A sourcebook of current theory, research and therapy. New York: Wiley; 1982. p. 1–18.

[2] Weiss RS. Loneliness: The experience of emotional and social isolation. Cambridge, MA: MIT Press; 1973.

[3] Perissinotto CM, Stijacic Cenzer I, Covinsky KE. Loneliness in older persons: a predictor of functional decline and death. Arch Intern Med. 2012;172(14):1078–83.

[4] Hawkley LC, Cacioppo JT. Loneliness matters: a theoretical and empirical review of consequences and mechanisms. Ann Behav Med. 2010;40(2):218–27.

[5] Cohen-Mansfield J, Hazan H, Lerman Y, Shalom V. Correlates and predictors of loneliness in older-adults: a review of quantitative results informed by qualitative insights. Int Psychogeriatr. 2016;28(4):557–76.

[6] Ong AD, Uchino BN, Wethington E. Loneliness and health in older adults: a mini-review and synthesis. Gerontology. 2016;62(4):443–9.

[7] Holt-Lunstad J, Smith TB, Baker M, Harris T, Stephenson D. Loneliness and social isolation as risk factors for mortality: a meta-analytic review. Perspect Psychol Sci. 2015;10(2):227–37.

[8] Valtorta NK, Kanaan M, Gilbody S, Ronzi S, Hanratty B. Loneliness and social isolation as risk factors for coronary heart disease and stroke: systematic review and meta-analysis of longitudinal observational studies. Heart. 2016;102(13):1009–16.

[9] Kuiper JS, Zuidersma M, Oude Voshaar RC, Zuidema SU, van den Heuvel ER, Stolk RP, et al. Social relationships and risk of dementia: A systematic review and meta-analysis of longitudinal cohort studies. Ageing Res Rev. 2015;22:39–57.

[10] Erzen E, Çikrikci Ö. The effect of loneliness on depression: A meta-analysis. Int J Soc Psychiatry. 2018;64(5):427–35.

[11] Calati R, Ferrari C, Brittner M, Oasi O, Olié E, Carvalho AF, et al. Suicidal thoughts and behaviors and social isolation: A narrative review of the literature. J Affect Disord. 2019;245:653–67.

[12] Leigh-Hunt N, Bagguley D, Bash K, Turner V, Turnbull S, Valtorta N, et al. An overview of systematic reviews on the public health consequences of social isolation and loneliness. Public Health. 2017;152:157–71.

[13] Fong T, Nourbakhsh I, Dautenhahn K. A survey of socially interactive robots. Rob Auton Syst. 2003;42(3-4):143–66.

[14] Breazeal C. Social robots for health applications. In: 2011 Annual International Conference of the IEEE Engineering in Medicine and Biology Society. IEEE; 2011. p. 5368–71.

[15] Robinson H, MacDonald B, Kerse N, Broadbent E. The psychosocial effects of a companion robot: a randomized controlled trial. J Am Med Dir Assoc. 2013;14(9):661–7.

[16] Moyle W, Jones C, Sung B, Bramble M, O’Dwyer S, Blumenstein M, et al. What effect does an animal robot called CuDDler have on the engagement and emotional response of older people with dementia? A pilot feasibility study. Int J Soc Robot. 2016;8(1):145–56.

[17] Pu L, Moyle W, Jones C, Todorovic M. The effectiveness of social robots for older adults: a systematic review and meta-analysis of randomized controlled studies. Gerontologist. 2019;59(1):e37–51.

[18] Abdi J, Al-Hindawi A, Ng T, Vizcaychipi MP. Scoping review on the use of socially assistive robot technology in elderly care. BMJ Open. 2018;8(2):e018815.

[19] Moher D, Liberati A, Tetzlaff J, Altman DG, PRISMA Group. Preferred reporting items for systematic reviews and meta-analyses: the PRISMA statement. PLoS Med. 2009;6(7):e1000097.

[20] Higgins JPT, Thomas J, Chandler J, Cumpston M, Li T, page MJ, et al., editors. Cochrane Handbook for Systematic Reviews of Interventions version 6.3 (updated February 2022). Cochrane; 2022. Available from www.training.cochrane.org/handbook.

[21] Russell D, Peplau LA, Cutrona CE. The revised UCLA Loneliness Scale: concurrent and discriminant validity evidence. J Pers Soc Psychol. 1980;39(3):472–80.

[22] De Jong Gierveld J, Van Tilburg T. A 6-item scale for overall, emotional, and social loneliness: confirmatory tests on survey data. Res Aging. 2006;28(5):582–98.

[23] Campaign to End Loneliness. Measuring your impact on loneliness in later life. London: Campaign to End Loneliness; 2015.

[24] Zimet GD, Dahlem NW, Zimet SG, Farley GK. The multidimensional scale of perceived social support. J Pers Assess. 1988;52(1):30–41.

[25] Lubben J, Blozik E, Gillmann G, Iliffe S, von Renteln Kruse W, Beck JC, et al. Performance of an abbreviated version of the Lubben Social Network Scale among three European community-dwelling older adult populations. Gerontologist. 2006;46(4):503–13.

[26] WHOQOL Group. Development of the World Health Organization WHOQOL-BREF quality of life assessment. Psychol Med. 1998;28(3):551–8.

[27] EuroQol Group. EuroQol--a new facility for the measurement of health-related quality of life. Health Policy. 1990;16(3):199–208.

[28] Yesavage JA, Brink TL, Rose TL, Lum O, Huang V, Adey M, et al. Development and validation of a geriatric depression screening scale: a preliminary report. J Psychiatr Res. 1982;17(1):37–49.

[29] Kroenke K, Spitzer RL, Williams JB. The PHQ-9: validity of a brief depression severity measure. J Gen Intern Med. 2001;16(9):606–13.

[30] Spitzer RL, Kroenke K, Williams JBW, Löwe B. A brief measure for assessing generalized anxiety disorder: the GAD-7. Arch Intern Med. 2006;166(10):1092–7.

[31] Spielberger CD, Gorsuch RL, Lushene R, Vagg PR, Jacobs GA. Manual for the State-Trait Anxiety Inventory. Palo Alto, CA: Consulting Psychologists Press; 1983.

[32] Folstein MF, Folstein SE, McHugh PR. “Mini-mental state”. A practical method for grading the cognitive state of patients for the clinician. J Psychiatr Res. 1975;12(3):189–98.

[33] Nasreddine ZS, Phillips NA, Bédirian V, Charbonneau S, Whitehead V, Collin I, et al. The Montreal Cognitive Assessment, MoCA: a brief screening tool for mild cognitive impairment. J Am Geriatr Soc. 2005;53(4):695–9.

[34] Veritas Health Innovation. Covidence systematic review software. Melbourne, Australia; Available at www.covidence.org.

[35] Sterne JAC, Savović J, page MJ, Elbers RG, Blencowe NS, Boutron I, et al. RoB 2: a revised tool for assessing risk of bias in randomised trials. BMJ. 2019;366:l4898.

[36] Sterne JA, Hernán MA, Reeves BC, Savović J, Berkman ND, Viswanathan M, et al. ROBINS-I: a tool for assessing risk of bias in non-randomised studies of interventions. BMJ. 2016;355:i4919.

[37] Critical Appraisal Skills Programme. CASP Qualitative Checklist. Available at: https://casp-uk.net/casp-tools-checklists/. Accessed: 2022 May 10.

[38] Higgins JPT, Thompson SG, Deeks JJ, Altman DG. Measuring inconsistency in meta-analyses. BMJ. 2003;327(7414):557–60.

[39] Guyatt GH, Oxman AD, Vist GE, Kunz R, Falck-Ytter Y, Alonso-Coello P, et al. GRADE: an emerging consensus on rating quality of evidence and strength of recommendations. BMJ. 2008;336(7650):924–6.

[40] Egger M, Davey Smith G, Schneider M, Minder C. Bias in meta-analysis detected by a simple, graphical test. BMJ. 1997;315(7109):629–34.

[41] Begg CB, Mazumdar M. Operating characteristics of a rank correlation test for publication bias. Biometrics. 1994;50(4):1088–101.

